# Temporal trends in COVID-19 outcomes among patients with systemic autoimmune rheumatic diseases: From the first wave to Omicron

**DOI:** 10.1101/2022.06.19.22276599

**Authors:** Yumeko Kawano, Naomi J. Patel, Xiaosong Wang, Claire E. Cook, Kathleen M.M. Vanni, Emily N. Kowalski, Emily P. Banasiak, Grace Qian, Michael DiIorio, Tiffany Y. T. Hsu, Michael E. Weinblatt, Derrick J. Todd, Zachary S. Wallace, Jeffrey A. Sparks

## Abstract

**Objectives:** To investigate temporal trends in incidence and severity of COVID-19 among patients with systemic autoimmune rheumatic diseases (SARDs) from the first wave through the Omicron wave.

**Methods:** We conducted a retrospective cohort study investigating COVID-19 outcomes among SARD patients systematically identified to have confirmed COVID-19 from March 1, 2020 to January 31, 2022 at a large healthcare system in Massachusetts. We tabulated COVID-19 counts of total and severe cases (hospitalizations or deaths) and compared the proportion with severe COVID-19 by calendar period and by vaccination status. We used logistic regression to estimate the ORs for severe COVID-19 for each period compared to the early COVID-19 period (reference group).

**Results:** We identified 1449 SARD patients with COVID-19 (mean age 58.4 years, 75.2% female, 33.9% rheumatoid arthritis). There were 399 (27.5%) cases of severe COVID-19. The proportion of severe COVID-19 outcomes declined over calendar time (p for trend <0.001); 45.6% of cases were severe in the early COVID-19 period (March 1-June 30, 2020) vs. 14.7% in the Omicron wave (December 17, 2021-January 31, 2022; adjusted odds ratio 0.29, 95%CI 0.19-0.43). A higher proportion of those unvaccinated were severe compared to not severe cases (78.4% vs. 59.5%).

**Conclusions:** The proportion of SARD patients with severe COVID-19 has diminished since early in the pandemic, particularly during the most recent time periods, including the Omicron wave. Advances in prevention, diagnosis, and treatment of COVID-19 may have improved outcomes among SARD patients.

**KEY MESSAGES:** *What is already known about this subject?:*

- Patients with systemic autoimmune rheumatic diseases (SARDs) may be at increased risk for severe COVID-19, defined as hospitalization or death.
- Previous studies of SARD patients suggested improving COVID-19 outcomes over calendar time, but most were performed prior to the wide availability of COVID-19 vaccines or the Omicron wave that was characterized by high infectivity.

*What does this study add?:*

- The proportion of SARD patients with severe COVID-19 outcomes was lower over calendar time
- The adjusted odds ratio of severe COVID-19 in the Omicron wave was 0.29 (95%CI 0.19-0.43) compared to early COVID-19 period.
- The absolute number of severe COVID-19 cases during the peak of the Omicron variant wave was similar to the peaks of other waves.
- SARD patients with severe vs. not severe COVID-19 were more likely to be unvaccinated.

*How might this impact on clinical practice or future developments?:*

- These findings suggest that advances in COVID-19 prevention, diagnosis, and treatment have contributed to improved outcomes among SARD patients over calendar time.
- Future studies should extend findings into future viral variants and consider the roles of waning immunity after vaccination or natural infection among SARD patients who may still be vulnerable to severe COVID-19.

## INTRODUCTION

In the two years since coronavirus disease 2019 (COVID-19) was recognized as a global pandemic, significant strides have been made in the testing, prevention, and treatment of COVID-19. The most recent wave of infections caused by the “Omicron’’ variant has been reported to cause less severe outcomes in the general population compared to prior waves ^1–4^. Multiple factors including vaccinations, prior infection, increased testing, and effective treatments as well as intrinsic features of the variant likely contribute to these observations. However, whether such improved outcomes have also been observed in people with systemic autoimmune rheumatic diseases (SARDs) remains unclear. Some people with SARDs have been found to have an increased risk of severe outcomes from COVID-19, such as hospitalization and death. This has been attributed to altered underlying immunity, immunosuppression contributing to blunted responses to both natural infection and vaccination, and end-organ damage from the SARD ^5 6^. The impact of vaccination, testing, and outpatient and inpatient treatments during the Omicron wave in the United States may have contributed to improved temporal COVID-19 outcomes among people with SARDs.

Previous studies have investigated temporal trends in COVID-19 outcomes among SARD patients prior to the Omicron wave. A Swedish study compared patients with inflammatory joint diseases to matched comparators and found worse outcomes, particularly early in the pandemic ^7^. A small cohort study in Ireland found no improvement in hospitalization or mortality rates in the first three waves of the pandemic ^8^. Two other studies performed about six months into the pandemic showed that excess risk of severe COVID-19 among SARD patients was similar to the general population ^9^ ^10^. It remains unclear whether outcomes have improved in recent time periods for patients with systemic autoimmune rheumatic diseases.

We aimed to investigate temporal trends in incidence and severity of COVID-19 among SARD patients. We hypothesized that the proportion of SARD patients experiencing severe COVID-19 has improved since early in the pandemic.

## METHODS

### Study design and population

We performed a retrospective cohort study investigating temporal trends of COVID-19 outcomes among SARD patients throughout the pandemic (from March 1, 2020 to January 31, 2022) at the Mass General Brigham (MGB) HealthCare system in the greater Boston, Massachusetts area. MGB is composed of 14 hospitals including Massachusetts General Hospital and Brigham and Women’s Hospital and affiliated community health centers. The study was approved by the MGB Institutional Review Board.

### Identification of COVID-19 cases and SARD patients

As previously described in more detail we systematically identified all SARD patients with PCR-confirmed SARS-CoV-2 infection using electronic query ^6^ ^10–14^. Our EHR also flags patients who had positive testing either at home (e.g., patients who notified their rheumatologist about a positive rapid antigen test through the secure patient portal) or outside of the MGB system (e.g., admitted patient with COVID-19 transferred from an outside hospital). These lists were filtered by the presence of at least one ICD-9 or ICD-10 code for SARD as a sensitive screen. This was further supplemented by direct referrals to our study team from rheumatologists who learned of patients’ positive tests during a clinical encounter. Among these lists, the presence of prevalent SARD and SARS-CoV-2 infection were confirmed by medical record review. As in our previous studies, we excluded participants only being treated for osteoarthritis, fibromyalgia, mechanical back pain, Raynaud’s phenomenon, gout, or pseudogout since these conditions are not typically treated with systemic immunomodulators and are often managed by non-rheumatologists ^6^ ^10–14^. Thus, all patients included had a confirmed SARD diagnosis with verified SARS-CoV-2 infection between March 1, 2020 and January 31, 2022 (**Supplementary Figure 1**).

### Exposure variable: Time periods throughout the pandemic

The date of COVID-19 onset was determined by the first date of SARS-CoV-2 test positivity for those with PCR tests in the MGB system, date of first COVID-19 flag, or date of positive test from medical record review or referral. We *a priori* divided calendar time into periods based on changes in viral epidemiology and care advances. The “Early COVID-19 period” was March 1 to June 30, 2020, corresponding to when Massachusetts experienced the first wave of COVID-19. The “Early treatment period” was July 1, 2020 to January 31, 2021, corresponding to seminal advances in treatment of hospitalized patients with dexamethasone and remdesivir as well as a wave of cases in the fall/winter ^15^ ^16^. The “Early vaccination” period was February 1 to June 30, 2021, corresponding to the initial rollout of COVID-19 vaccines to high-risk populations starting on February 1, 2021 ^17–19^. The “Additional vaccination and Delta wave” period was July 1 to December 16, 2022, corresponding to recommendations for additional doses of vaccines to immunocompromised patients as well as a fall surge of cases due to the Delta variant ^20^. The “Omicron wave” period was December 17, 2021 to January 31, 2022, corresponding to the large local surge of cases due to the Omicron variant.

### Outcome variables: Severe COVID-19

As in previous studies, we defined severe COVID-19 as a composite of hospitalization or death within 30 days from COVID-19 date ^5^. Medical record review was performed to determine these outcomes and to identify patients who received mechanical ventilation. This was supplemented by electronic query and information from the referring rheumatologist for those who were hospitalized outside the MGB system. We also examined the individual outcomes of hospitalization, mechanical ventilation, and death from COVID-19.

### Other characteristics

We collected information on demographics, comorbidities, vaccination status, and rheumatic disease characteristics and medications. Age, sex, race (White, Black, Asian/Hawaiian/Pacific Islander, or Other/unknown) and Hispanic ethnicity were identified from electronic query. Comorbidities were collected from medical record review and included hypertension, diabetes mellitus, obesity, cardiovascular disease, obstructive lung disease (including asthma, chronic obstructive pulmonary disease, and obstructive sleep apnea), and interstitial lung disease. We classified vaccination status as follows: 1) unvaccinated or pre-vaccine, 2) partially vaccinated (occurring between Day 0 to Day 13 of 2-dose mRNA vaccines or between Day 0 to Day 13 of the 1-dose Johnson & Johnson-Janssen [J&J] vaccine), 3) two doses mRNA or one dose J&J (14 days after completion of the initial series) and 4) additional doses (Day 14 or later after third vaccine dose for those who initially received 2-dose mRNA vaccines or Day 14 or later after second vaccine dose for those who initially received 1-dose J&J vaccine per CDC recommendations for an additional dose)^21^. (**Supplementary Figure 2**).

SARD characteristics were obtained from medical record review. These included specific type of SARD, glucocorticoid use/dose (none, low dose [1-10mg/day of prednisone-equivalent], moderate/high dose (>10 mg), and unknown dose), disease-modifying antirheumatic drug (DMARD) use (categorized by specific drug for conventional synthetic DMARDs or mechanism of action for biologic DMARDs) at time of COVID-19 diagnosis, and preceding SARD activity per review of notes from the medical record (categorized as remission/low, moderate/severe, or unknown).

Treatment of COVID-19 was also collected by medical record review and supplemented by electronic query: neutralizing monoclonal antibodies (typically given as outpatient for COVID-19 treatment rather than inpatient or as post-exposure prophylaxis during our study period), remdesivir (only used as inpatient during our study period), convalescent plasma, dexamethasone (or other high-dose glucocorticoids used inpatient to treat COVID-19), tocilizumab (to treat inpatient COVID-19 rather than underlying SARD), baricitinib (used inpatient to treat COVID-19 rather than underlying SARD) ^22^. No SARD patients were documented to have received nirmatrelvir/ritonavir (Paxlovid) or molnupiravir for COVID-19 treatment during the study period ^23^.

### Statistical analysis

We graphed weekly counts of total COVID-19 cases and severe outcomes among SARD patients throughout the entire study period. We reported frequencies and proportions of demographics, comorbidities, vaccination status, type of COVID-19 test, and SARD characteristics overall and stratified by the five calendar periods. Since severe COVID-19 during the Omicron wave period has been reported to be relatively uncommon in the general population,^1^ we also reported on characteristics of SARD patients with severe vs. not severe COVID-19 during the period. We also reported a case series of the SARD patients who died of COVID-19 during the Omicron wave period. We determined whether each case was due to underlying SARD/immunosuppression or due to other comorbidities from consensus discussion from the study rheumatologists (YK, NJP, ZSW, and JAS).

We reported the proportion of severe COVID-19 outcomes that occurred during each calendar period and calculated a p for trend across the five calendar periods. We also stratified cases by severe vs. not severe and calculated the proportion in each category by vaccination status. We performed logistic regression to estimate the odds ratios (ORs) and 95% confidence intervals (CIs) for severe COVID-19 for each period compared to the early pandemic period as the reference. The base model was unadjusted. The multivariable model adjusted for age, sex, and race. We considered a two-sided p value of <0.05 as statistically significant. SAS v9.4 (Cary, NC) was used for all analyses.

## RESULTS

### COVID-19 cases among SARD patients

From March 1, 2020 to January 31, 2022, we identified 1,449 SARD patients with confirmed COVID-19 (see flow diagram for the analyzed sample in **Supplemental Figure 1**). **Figure 1** shows the weekly counts of total and severe COVID-19 cases. The tallest peak of cases occurred during the Omicron Wave period. There were 261, 492, 123, 172, and 401 total cases in the Early COVID-19, Early treatment, Early vaccination, Additional vaccination and Delta wave, and Omicron wave periods, respectively.

**Figure 1:**
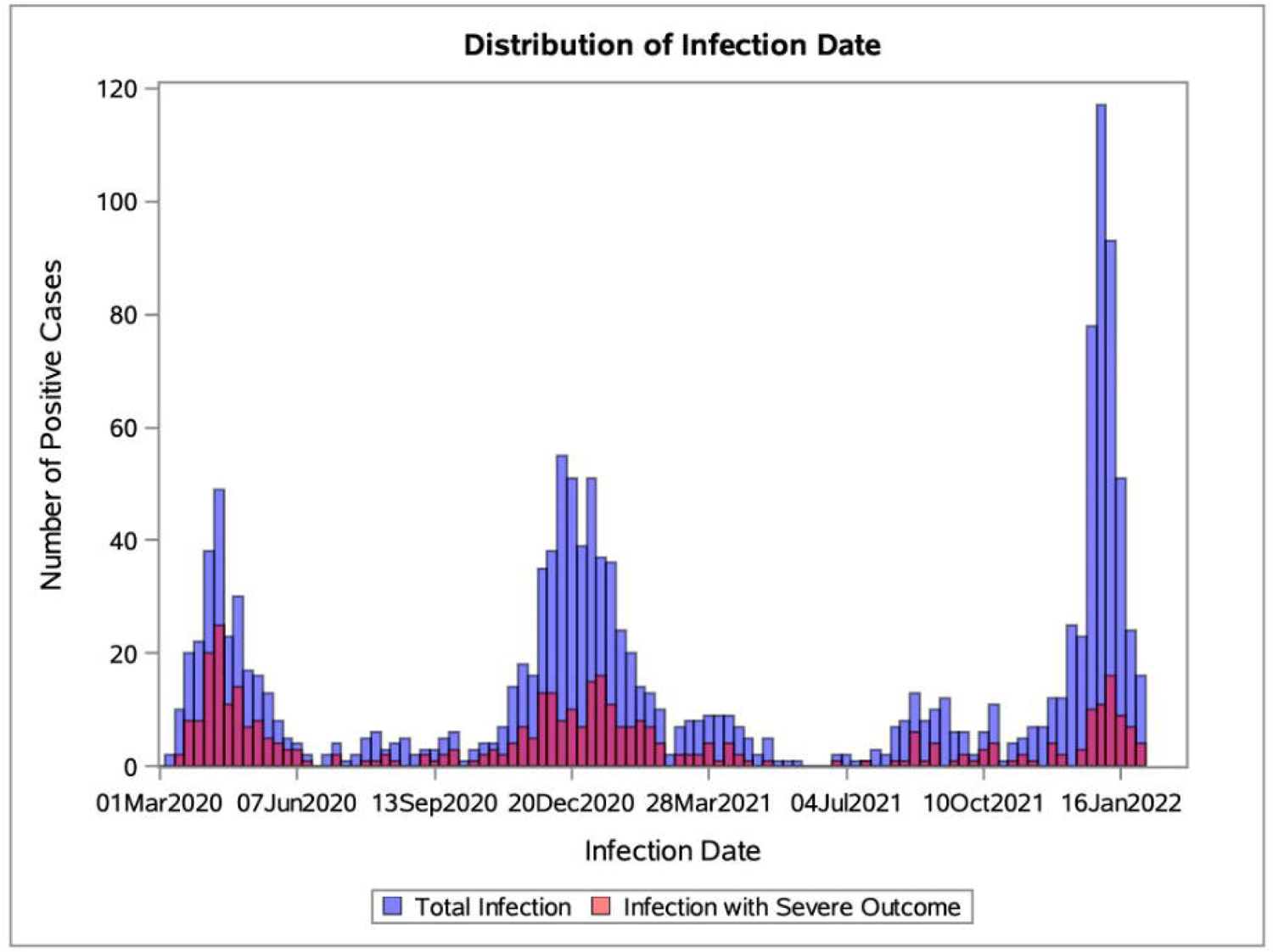
Total and severe COVID-19 case counts over time. Case counts of SARS-CoV-2 infections over calendar time with total infections shown in blue and infections with severe outcomes in red.

### Characteristics of SARD patients with COVID-19

**Table 1** shows the demographics, comorbidities, SARS-CoV-2 testing type, and COVID-19 vaccination status. Mean age of the entire study sample was 58.4 years (SD 17.5). Cases in the Early COVID-19 period tended to be older compared to cases in the Omicron wave period (mean age 63.1 years vs. 54.2). The overall sample was 71.4% White, 11.2% Black, and 3.6% Asian/Hawaiian/Pacific Islander. The most common comorbidity was hypertension (43.8%). The vaccination status for the entire sample at the time of infection was 64.7% prevaccination/unvaccinated, 3.5% partially vaccinated, 15.7% 2 doses mRNA or 1 dose J&J, and 16.1% additional doses. Breakthrough infections were particularly common in the Omicron wave: 136 cases (33.8% of cases in the period) occurred among those who received 2 doses mRNA or 1 dose J&J and 205 cases (51.1% of cases in this period) occurred among those with additional vaccine doses. Most cases were diagnosed with PCR testing. The Omicron wave was the only period where a substantial portion of cases were diagnosed with home rapid antigen testing (122/401 [30.4%] during this period).

**Table 1:** Demographics and clinical characteristics of rheumatic disease patients with COVID-19

|  | Time period |  |  |  |  |  |
| --- | --- | --- | --- | --- | --- | --- |
|  |  | Early COVID-19 | Early treatment | Early vaccination | Additional vaccination and Delta wave | Omicron wave |
|  | Overall (n = 1449) | March 1 2020 - June 30 2020 (n = 261) | July 1 2020 - Jan 31 2021 (n = 492) | Feb 1 2021 - June 30 2021 (n = 123) | July 1 2021 - Dec 16 2021 (n = 172) | Dec 17 2021 - Jan 31 2022 (n = 401) |
| **Age, years (mean ± SD)** | 58.4 (17.5) | 63.1 (16.6) | 59.3 (17.0) | 55.5 (16.9) | 56.8 (17.7) | 54.2 (17.6) |
| **Female sex, n (%)** | 1090 (75.2%) | 196 (75.1%) | 370 (75.2%) | 90 (73.2%) | 111 (64.5%) | 323 (80.5%) |
| **Race, n (%)** |  |  |  |  |  |  |
| White | 1035 (71.4%) | 154 (59.0%) | 353 (71.7%) | 93 (75.6%) | 136 (79.1%) | 299 (74.6%) |
| Black | 162 (11.2%) | 52 (19.9%) | 50 (10.2%) | 15 (12.2%) | 11 (6.4%) | 34 (8.5%) |
| Asian, Hawaiian or Pacific Islander | 52 (3.6%) | 9 (3.4%) | 19 (3.9%) | 6 (4.9%) | 7 (4.1%) | 11 (2.7%) |
| Other or unknown | 200 (13.8%) | 46 (17.6%) | 70 (14.2%) | 9 (7.3%) | 18 (10.5%) | 57 (14.2%) |
| **Hispanic ethnicity, n (%)** | 67 (4.6%) | 13 (5.0%) | 31 (6.3%) | 5 (4.1%) | 4 (2.3%) | 14 (3.5%) |
| **Comorbidities, n (%)** |  |  |  |  |  |  |
| Hypertension | 635 (43.8%) | 154 (59.0%) | 220 (44.7%) | 49 (39.8%) | 63 (36.6%) | 149 (37.2%) |
| Diabetes Mellitus | 234 (16.1%) | 61 (23.4%) | 92 (18.7%) | 20 (16.3%) | 20 (11.6%) | 41 (10.2%) |
| Obesity | 427 (29.5%) | 92 (35.2%) | 156 (31.7%) | 49 (39.8%) | 35 (20.3%) | 95 (23.7%) |
| Cardiovascular disease | 210 (14.5%) | 62 (23.8%) | 72 (14.6%) | 15 (12.2%) | 20 (11.6%) | 41 (10.2%) |
| Obstructive lung disease | 310 (21.4%) | 73 (28.0%) | 109 (22.2%) | 25 (20.3%) | 31 (18.0%) | 72 (18.0%) |
| Interstitial lung disease | 81 (6.0%) | 17 (6.5%) | 25 (5.1%) | 8 (6.5%) | 9 (5.2%) | 22 (5.5%) |
| **Vaccination status, n (%)** |  |  |  |  |  |  |
| Unvaccinated or pre-vaccine | 938 (64.7%) | 261 (100%) | 492 (100%) | 102 (82.9%) | 40 (23.3%) | 43 (10.7%) |
| Partially vaccinated | 50 (3.5%) | 0 (0%) | 0 (0%) | 19 (15.4%) | 14 (8.1%) | 17 (4.2%) |
| Two doses mRNA or one dose J&J | 228 (15.7%) | 0 (0%) | 0 (0%) | 2 (1.6%) | 90 (52.3%) | 136 (33.9%) |
| Additional doses | 233 (16.1%) | 0 (0%) | 0 (0%) | 0 (0%) | 28 (16.3%) | 205 (51.1%) |
| **SARS-CoV2 diagnosis method, n (%)** |  |  |  |  |  |  |
| PCR | 1126 (77.7%) | 246 (94.3%) | 426 (86.6%) | 100 (81.3%) | 129 (75.0%) | 225 (56.1%) |
| Antigen/rapid test | 123 (8.5%) | 0 (0%) | 0 (0%) | 0 (0%) | 1 (0.6%) | 122 (30.4%) |
| Other or unknown | 200 (13.8%) | 15 (5.7%) | 66 (13.4%) | 23 (18.7%) | 42 (24.4%) | 54 (13.5%) |

COVID-19, coronavirus disease 2019; SD, standard deviation; mRNA, messenger ribonucleic acid;
J&J, Johnson and Johnson / Janssen; SARS-CoV-2, severe acute respiratory syndrome coronavirus
2; PCR, polymerase chain reaction

**Table 2** shows the SARD characteristics for the COVID-19 cases, stratified by the five calendar periods. The most common type of SARD was rheumatoid arthritis (33.9%), followed by psoriatic arthritis and spondyloarthritis (14.7%) and systemic lupus erythematosus (13.1%). Most patients (74.1%) were in remission or had low disease activity at time of COVID-19 onset. Baseline glucocorticoids were used in 26.0% of cases. The most commonly used DMARD was methotrexate (22.0%) followed by antimalarials (21.7%) and TNF inhibitors (20.2%). Rituximab was used in 9.2% of cases.

**Table 2:**
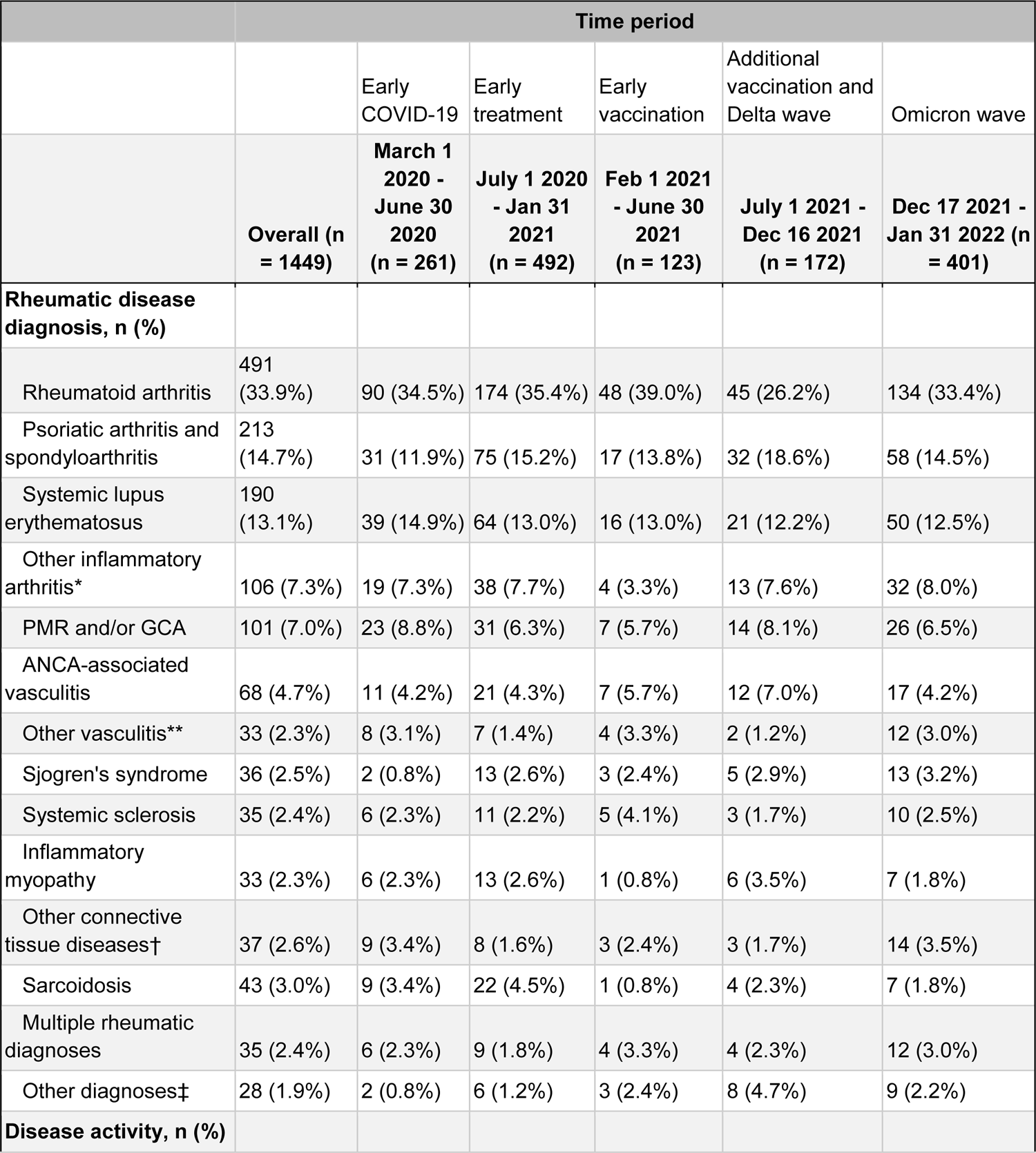
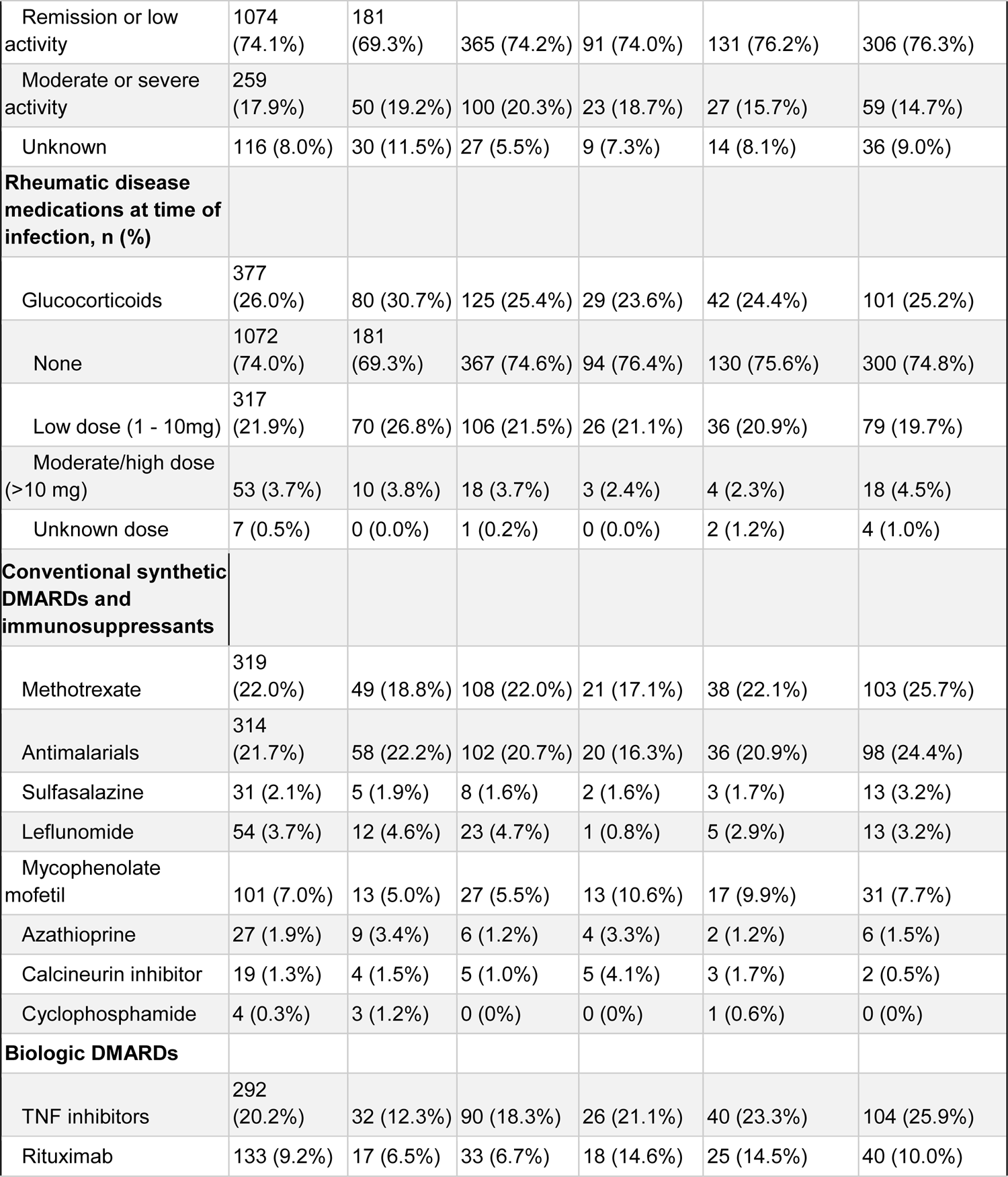
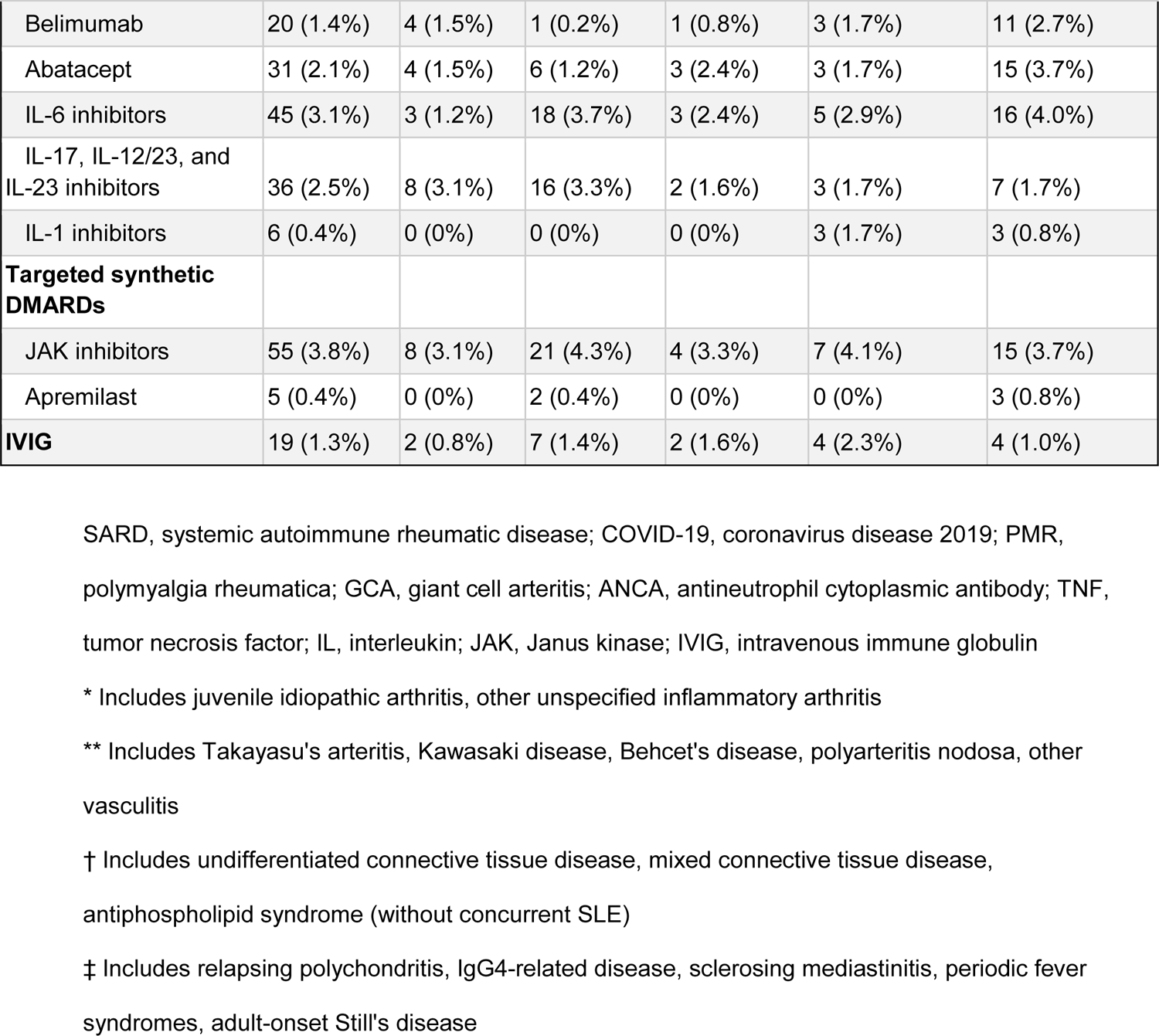
Rheumatic disease characteristics of SARD patients with COVID-19 over time

### Treatment and severe COVID-19 outcomes

The most common treatments for COVID-19 were remdesivir (14.2%), dexamethasone (13.2%), and neutralizing monoclonal antibodies (12.6%). Few SARD patients received tocilizumab (1.0%), baricitinib (0.2%), or convalescent plasma (0.5%) to treat COVID-19.

There were 399 (27.5%) who had the composite outcome of severe COVID-19 of hospitalization or death (**Table 3**). There were 391 (27.0%) hospitalizations and 60 (4.1%) deaths. The proportion of SARD patients experiencing severe COVID-19 decreased over the calendar periods. The total and proportion of severe COVID-19 in each calendar period was 119 (45.6%), 144 (29.3%), 41 (33.3%), 36 (20.9%), and 59 (14.7%) in the Early COVID-19, Early treatment, Early vaccination, Additional vaccination and Delta wave, and Omicron wave periods, respectively (**Figure 2**, p for trend <0.001). Compared to the reference of the Early COVID-19 period, the multivariable ORs and 95%CIs for severe COVID-19 were 0.58 (0.41 to 0.81) in the Early treatment period, 0.89 (0.54 to 1.46) in the Early vaccination period, 0.39 (0.24 to 0.62) in the Additional vaccination and Delta wave period, and 0.29 (0.19 to 0.43) in the Omicron wave period, adjusted for age, sex, and race.

**Figure 2:**
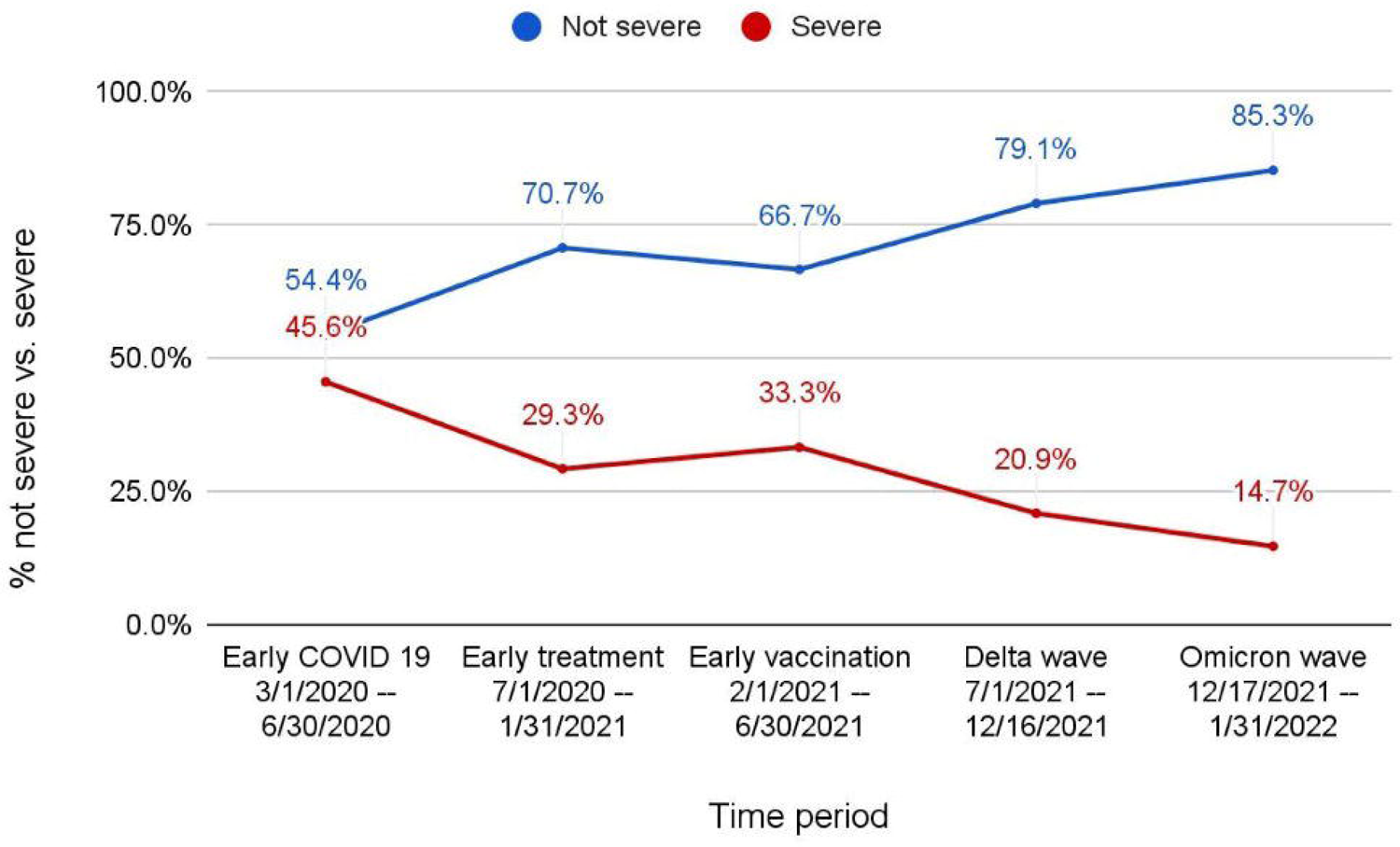
Proportion of cases with severe and non-severe COVID-19 in each time period. P value for trend.

**Table 3:**
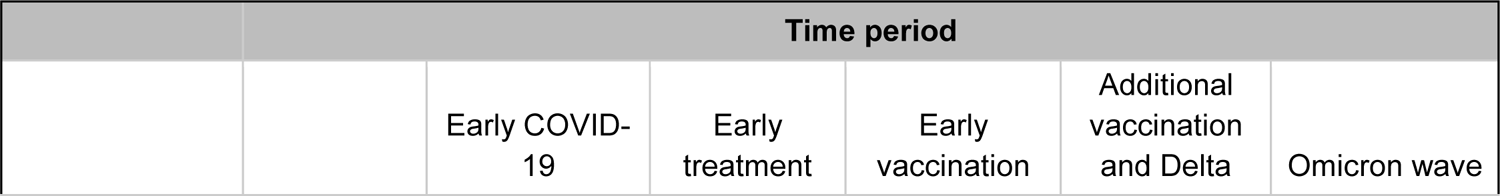
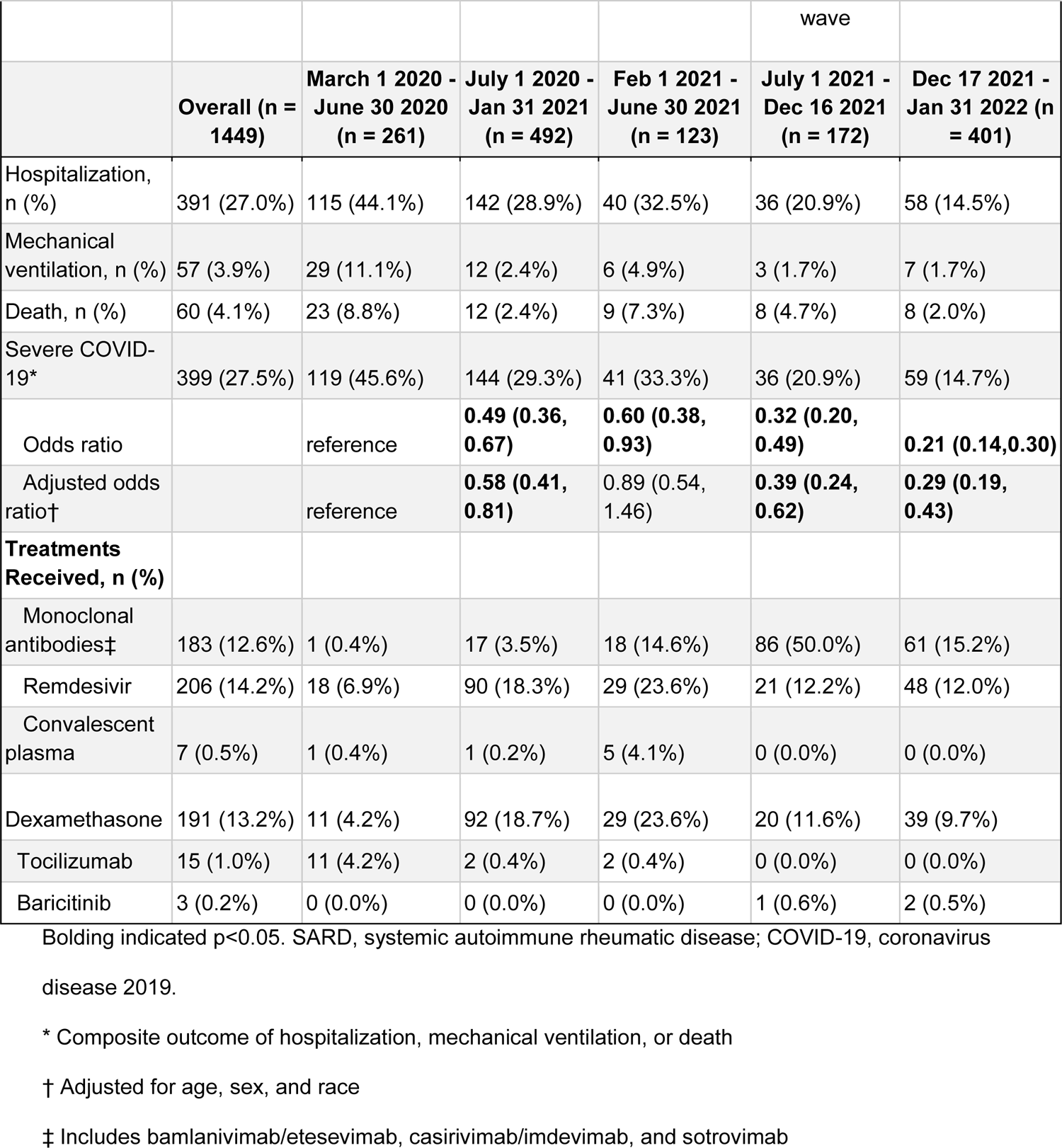
Outcomes and treatments of SARD patients over time

**Figure 3** displays the vaccination status stratified by severe or not severe COVID-19. Among the 399 severe cases, 78.4% were unvaccinated. Among the 1050 cases that were not severe, 59.5% were unvaccinated.

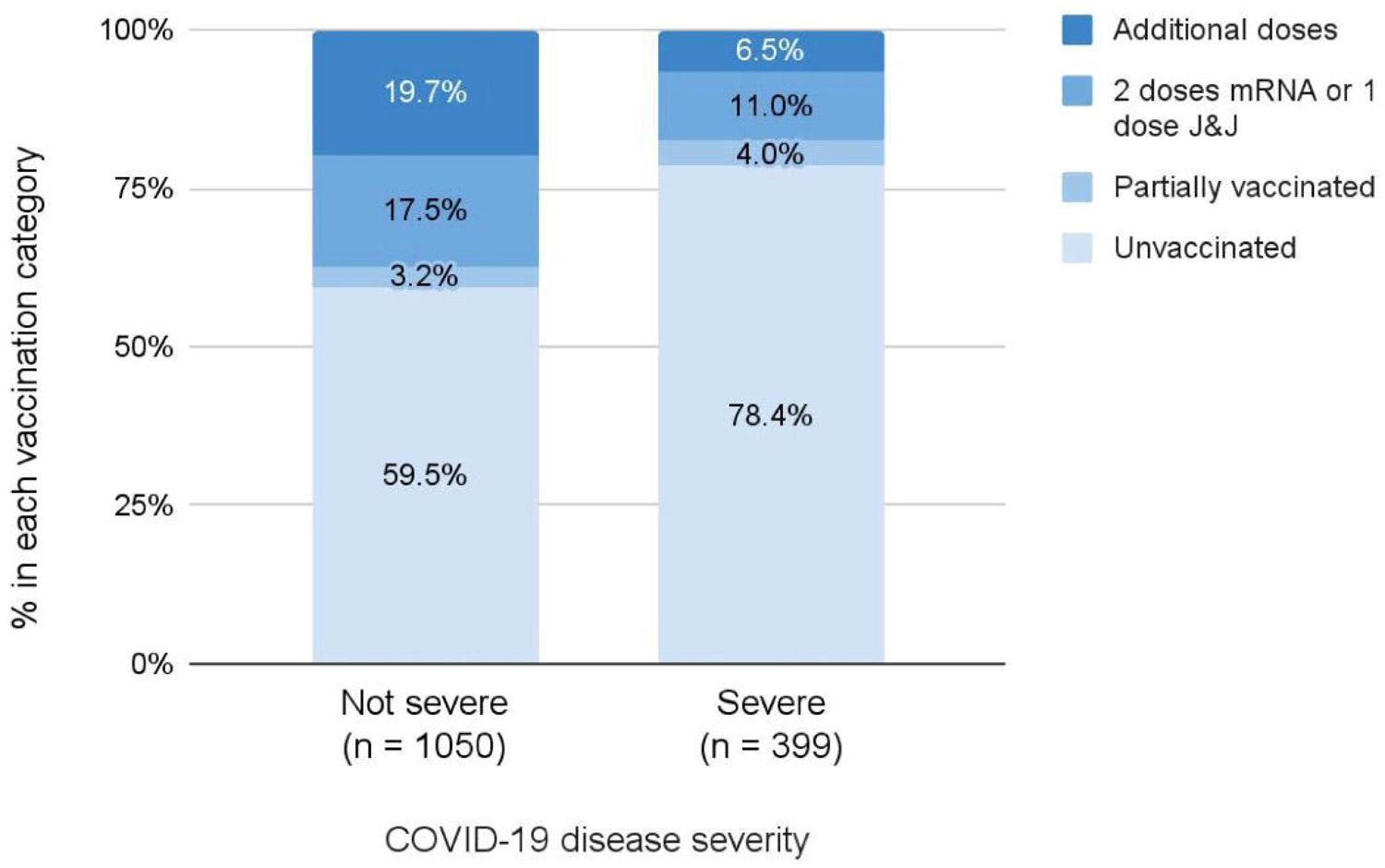

#### Severe COVID-19 outcomes during the Omicron wave period

**Supplemental Table 1** shows characteristics of the 59 SARD patients in the Omicron wave with severe COVID-19. Mean age was 66.9 years (SD 19.1), 76.3% were female, 10.2% had interstitial lung disease, 37.3% had rheumatoid arthritis, 47.5% were on glucocorticoids, 18.6% were on methotrexate, and 15.3% were on rituximab.

**Supplemental Table 2** shows the case series of the 8 SARD patients who died during the Omicron wave. Three of the deaths were likely due to underlying SARD / immunosuppression (case 1 on rituximab to treat ANCA-associated vasculitis; case 2 on abatacept to treat rheumatoid arthritis; case 3 on mycophenolic acid, tacrolimus, and prednisone for recent kidney transplant for ANCA-associated vasculitis). Two of the Omicron wave deaths were possibly due to underlying SARD / immunosuppression; the remaining three Omicron wave deaths were likely due to other comorbidities.

## DISCUSSION

In this large cohort study, we found that outcomes of COVID-19 in patients with SARDs have improved since the beginning of the pandemic. In particular, SARS-CoV-2 infections in the Omicron era were associated with a 71% reduction in the risk of hospitalization or death compared with the earliest time period. Despite these improvements, the absolute number of cases of severe COVID-19 was similar to that observed in other waves, suggesting that despite a reduced risk of severe disease, the Omicron wave had a substantial impact on patients with SARDs and the healthcare systems caring for them. The temporal improvement in outcomes is likely multifactorial, including differences in availability of testing, vaccination and other preventative strategies, hospital capacity, availability of effective treatments, depletion of susceptible individuals, and virulence of SARS-CoV2 variants.

Previous studies conducted earlier in the pandemic found improvement in hospitalization, mechanical ventilation, death, and other outcomes for patients with SARDs over time, even during the first 6 months of the pandemic ^9^ ^10^. However, no other study to date has examined the temporal trends in COVID-19 outcomes among SARDs extending up to the most recent Omicron wave. These findings are particularly relevant given the blunted vaccine response and associated higher risk of breakthrough infections that have been observed in some patients with SARDs ^24^.

The introduction of effective vaccines represented a key turning point in the pandemic and despite waning efficacy with time and against novel variants, they continue to provide important protection against severe disease ^25^. In our study of patients with SARDs, the majority of whom were on immunosuppressive treatments previously associated with blunted vaccine responses, we found that vaccination was associated with less severe COVID-19. These findings suggest that while some SARD patients on immunosuppressives may be at higher risk for breakthrough infection, vaccination provided important benefits for many of these patients if infected with SARS-CoV-2. Additional studies are needed to further evaluate the efficacy of COVID-19 vaccinations among patients with SARDs during the most recent Omicron wave, a time characterized by substantial waning efficacy against breakthrough infection in the general population ^26^ ^27^.

Despite temporal improvements in the risk of severe COVID-19, some patients with SARDs during the Omicron wave experienced hospitalization or death. Notably, these patients tended to have other comorbidities known to be associated with severe disease (e.g., interstitial lung disease, malignancy) and to be on treatments associated with substantially blunted immune responses to both vaccine and infection (e.g., B cell depletion) ^12^ ^28–30^. These findings highlight the need for ongoing risk mitigating strategies for many SARDs patients on such treatments as well as those with other comorbidities that may be related to their SARD (e.g., interstitial lung disease) or its treatment (e.g., cardiovascular comorbidities). In addition to shielding practices such as masking social distancing and avoiding indoor congregation, the recent introduction of pre-exposure prophylaxis with tixagevimab/cilgavimab, a monoclonal antibody against SARS-CoV-2, represents an important strategy for protecting our highest risk patients. However, tixagevimab/cilgavimab was studied in high risk, unvaccinated patients, the vast majority of whom did not have SARDs or similar conditions ^31^. Real world effectiveness studies of tixagevimab/cilgavimab are now being reported and will be informative for guiding ongoing risk mitigating strategies for patients with SARDs ^31^.

In addition to temporal improvements in the outcomes of COVID-19 among patients with SARDs during the ongoing pandemic, we also found other notable trends. First, there were shifts in the demographics of patients with COVID-19 during the course of the pandemic. For instance, there was a decrease in the proportion of SARDs patients with COVID-19 who identify as Black or Hispanic during the study period. There was also a decrease in the age of patients during the study period. These shifts are likely multifactorial, reflecting depletion of susceptible patients, changes in access to diagnostics and treatments, rates of vaccination, and other factors. Second, a large portion of infections are now diagnosed at home using rapid antigen tests. This shift in diagnostics will make it increasingly difficult to capture more mild infections for the purpose of epidemiologic studies like this one in both the general population and among SARDs. Leveraging electronic health record data, as we did here, will be an important way to capture and include patients like these in future studies.

Strengths of our study include the systematic identification of patients with SARDs in a large healthcare system that includes both tertiary care hospitals as well as community hospitals and their affiliated outpatient clinics. In contrast to studies relying only on administrative claims data, we were also able to capture COVID-19 diagnoses made using home rapid antigen testing because of the way these are captured in our electronic health record when reported to providers in our healthcare system. Additionally, we were able to confirm the SARD diagnosis and treatment, vaccination status at the time of infection, and details surrounding deaths and other outcomes with manual chart review.

Despite these strengths, our study has certain limitations. First, although we found temporal improvements in the outcomes of COVID-19 among patients with SARDs and an association between vaccination status and less severe disease, we cannot establish the causal effects of vaccinations, other risk mitigating strategies, and COVID-19 treatments on these improvements. Second, this study was conducted in a single healthcare system in Massachusetts, U.S. Our findings may not be generalizable to other areas of the U.S. or world because of differences in demographics as well as access to testing and treatment. Third, although we systematically identified cases in our healthcare system, patients who received COVID-19 diagnoses outside of our system may not have been captured, severe events occurring after diagnosis may have been missed, and patients with asymptomatic disease or mild courses may have been less likely to have testing or report their positive test. However, those with SARDs, especially those on immunosuppression, are likely to contact their clinicians in the context of a COVID-19 infection to receive guidance regarding the management of their SARD treatments and to obtain treatments.

In conclusion, there have been substantial improvements in the outcomes associated with COVID-19 among patients with SARDs since the pandemic began in March 2020. In particular, the Omicron wave was associated with the largest number of cases but the lowest risk of severe disease. Despite these findings, some patients with SARDs continue to experience severe disease, especially those on immunosuppressives known to blunt the response to vaccine and infection, as well as those with other severe comorbidities. Additional studies are needed to refine risk mitigating strategies for patients at highest risk for severe outcomes.

## Data Availability

All data produced in the present study are available upon reasonable request to the authors

---

**Supplemental Figure 1:**
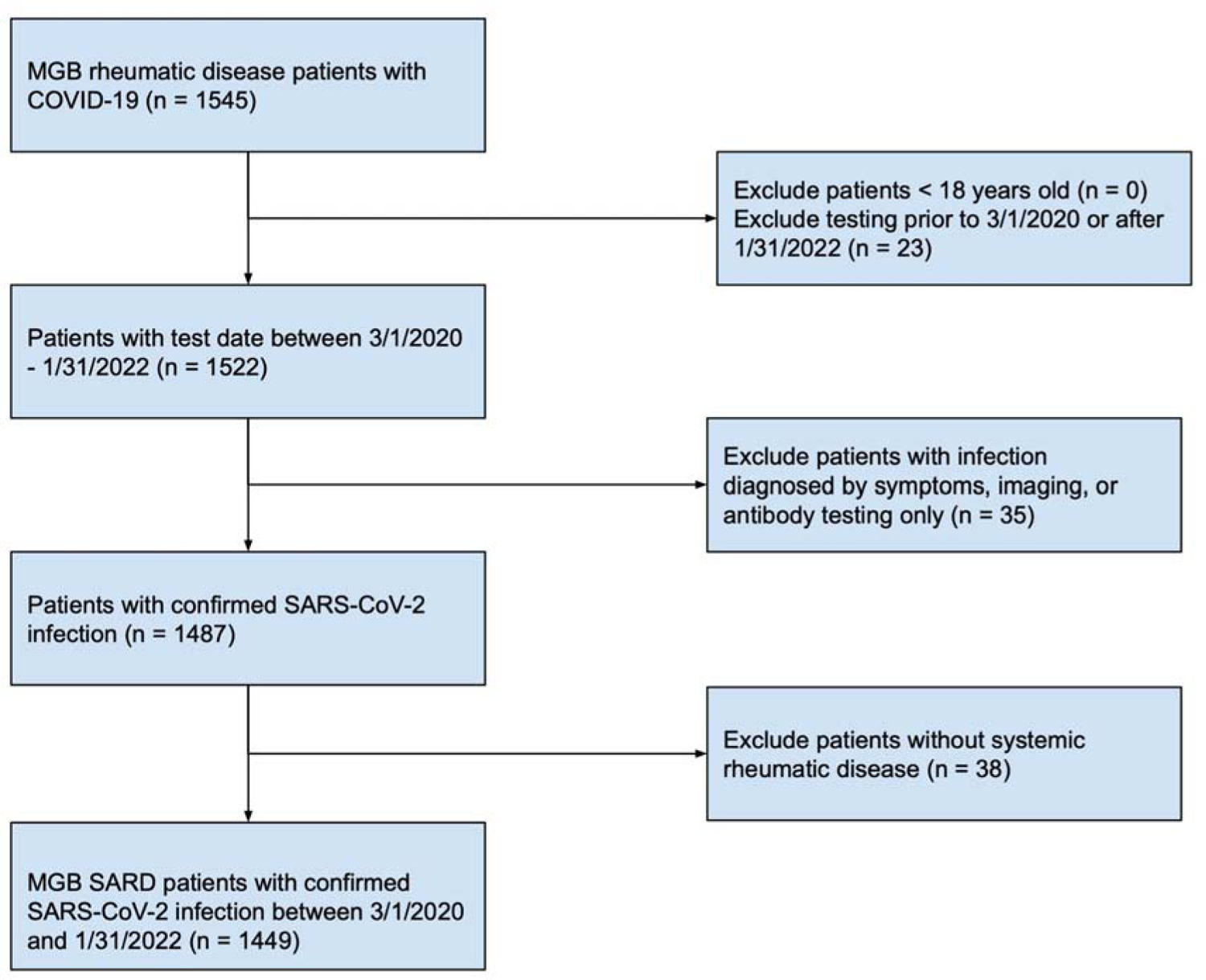
Flow chart of patient cohort selection. MGB, Mass General Brigham; SARS-CoV-2, severe acute respiratory syndrome coronavirus 2; SARD, systemic autoimmune rheumatic disease.

**Supplemental Figure 2:**
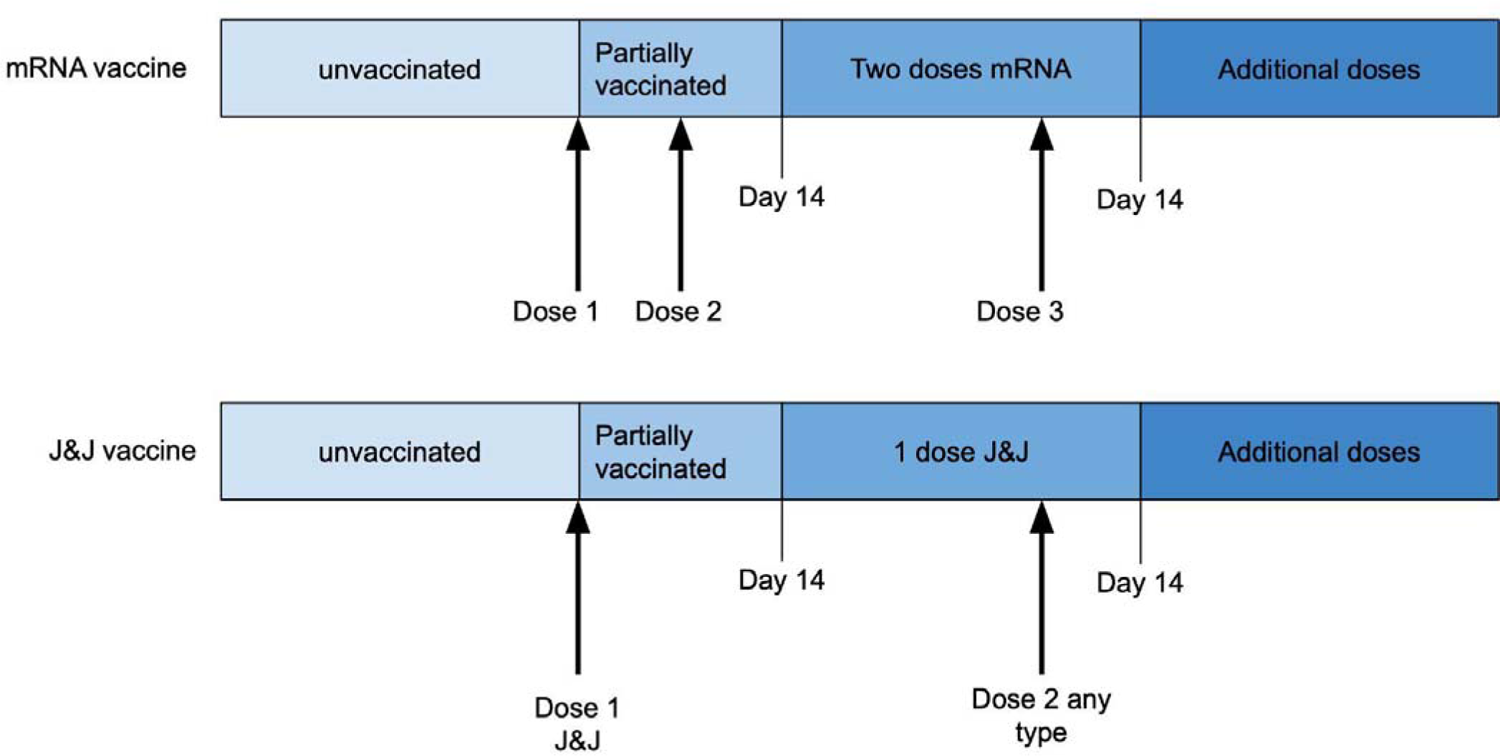
Definition of vaccination categories: “unvaccinated,” “partially vaccinated,” “two doses mRNA or 1 dose adenovirus,” and “additional doses.”

**Supplemental Table 1:**
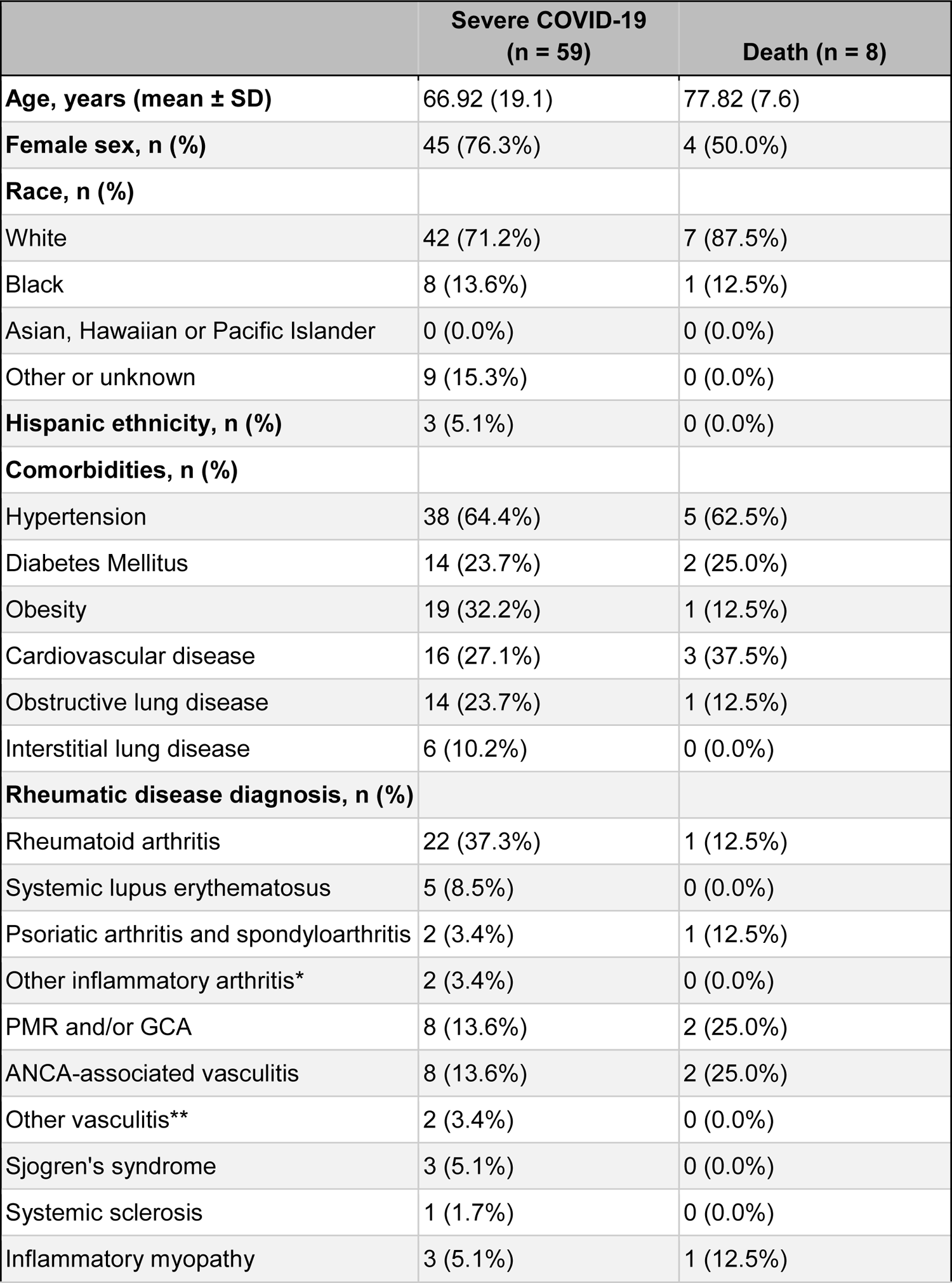
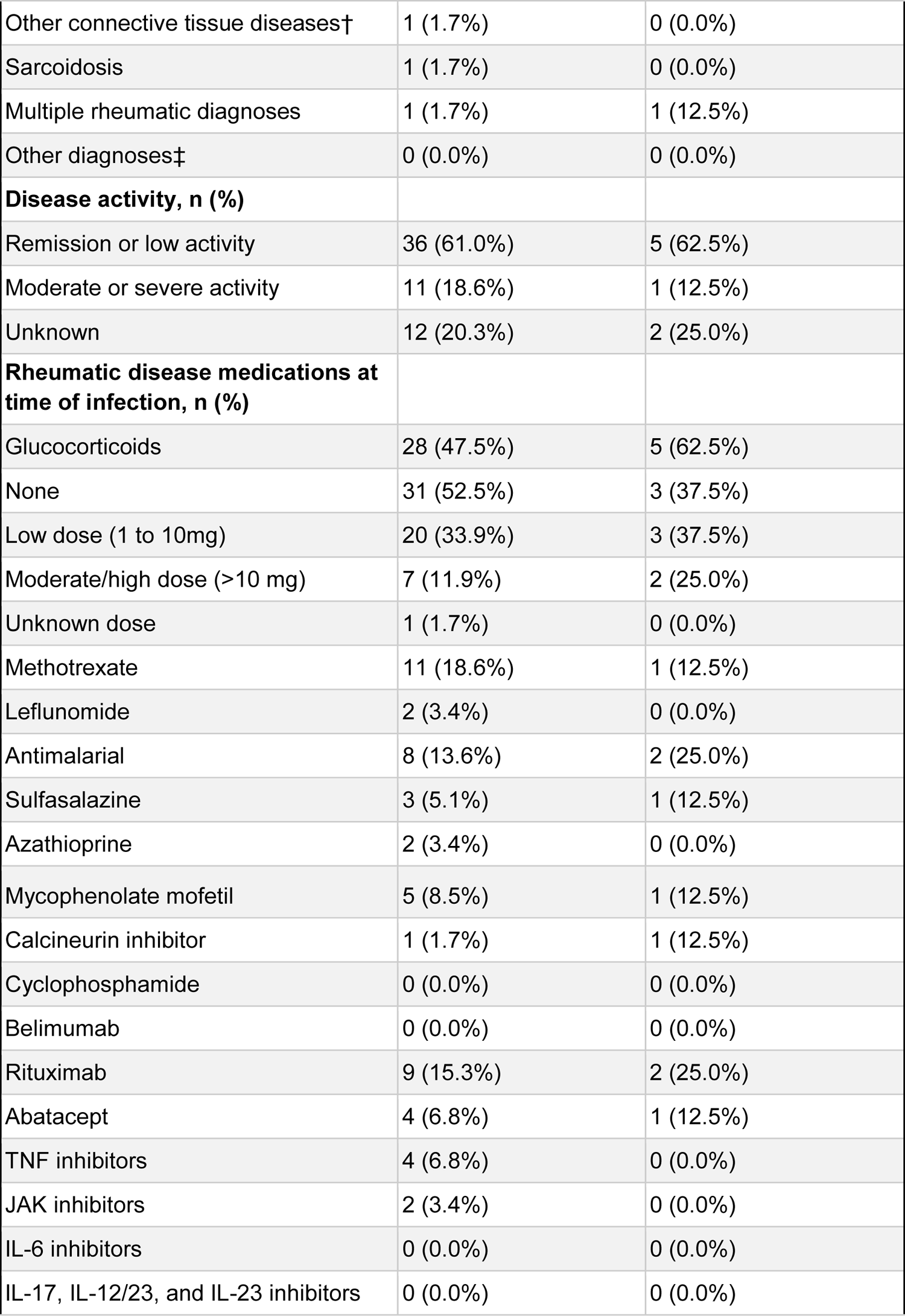
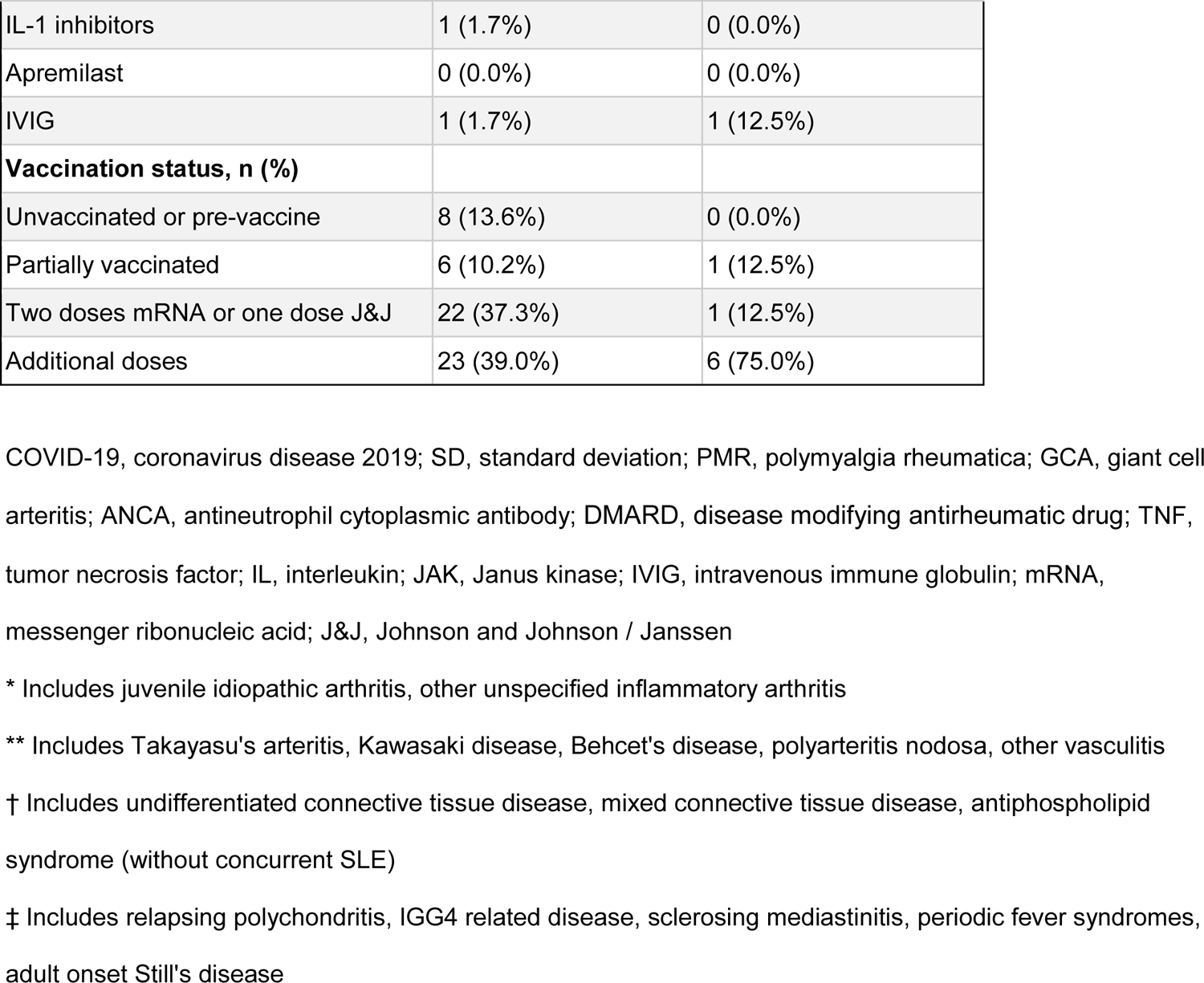
Characteristics of patients with severe COVID-19 during Omicron wave

**Supplemental Table 2:**
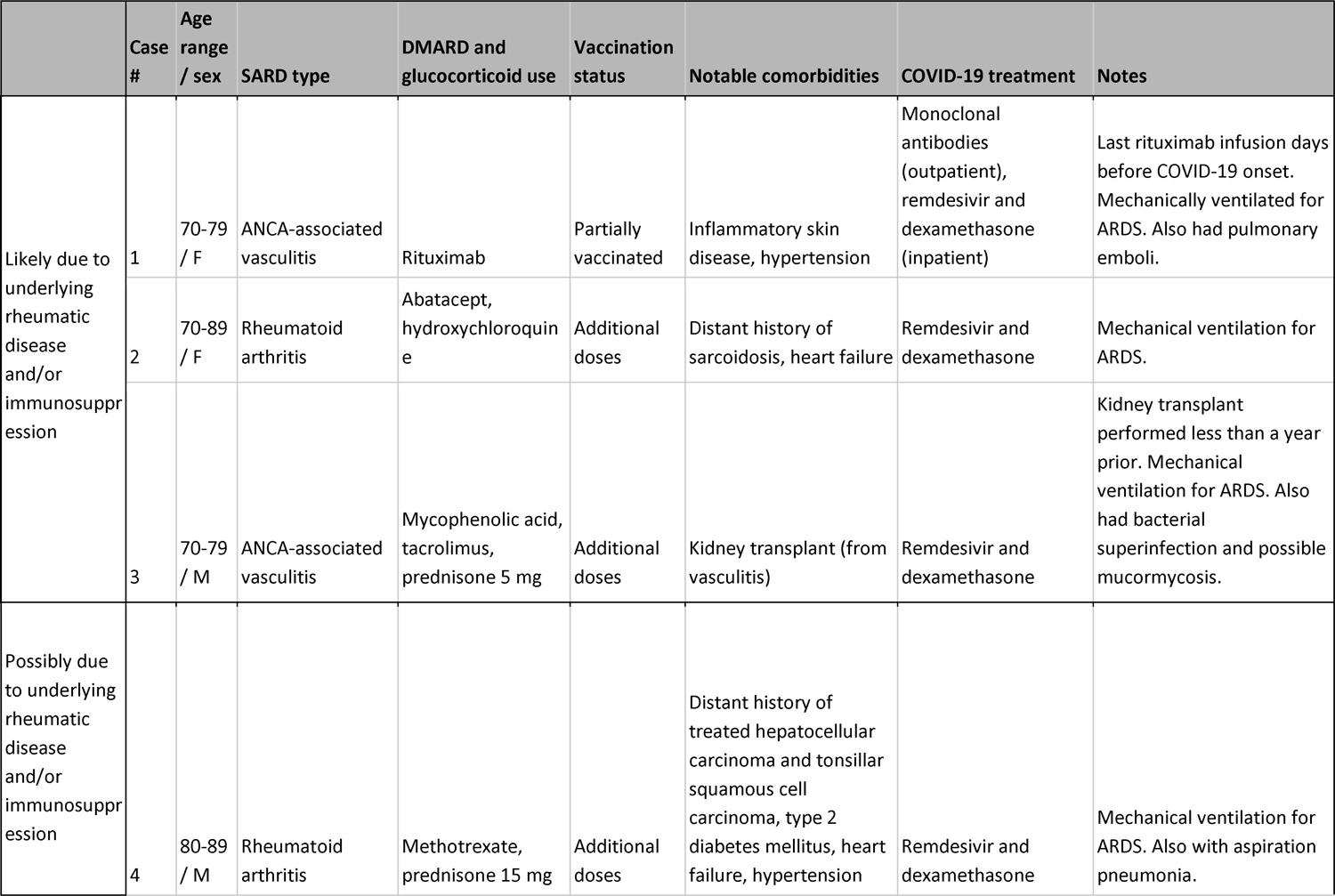
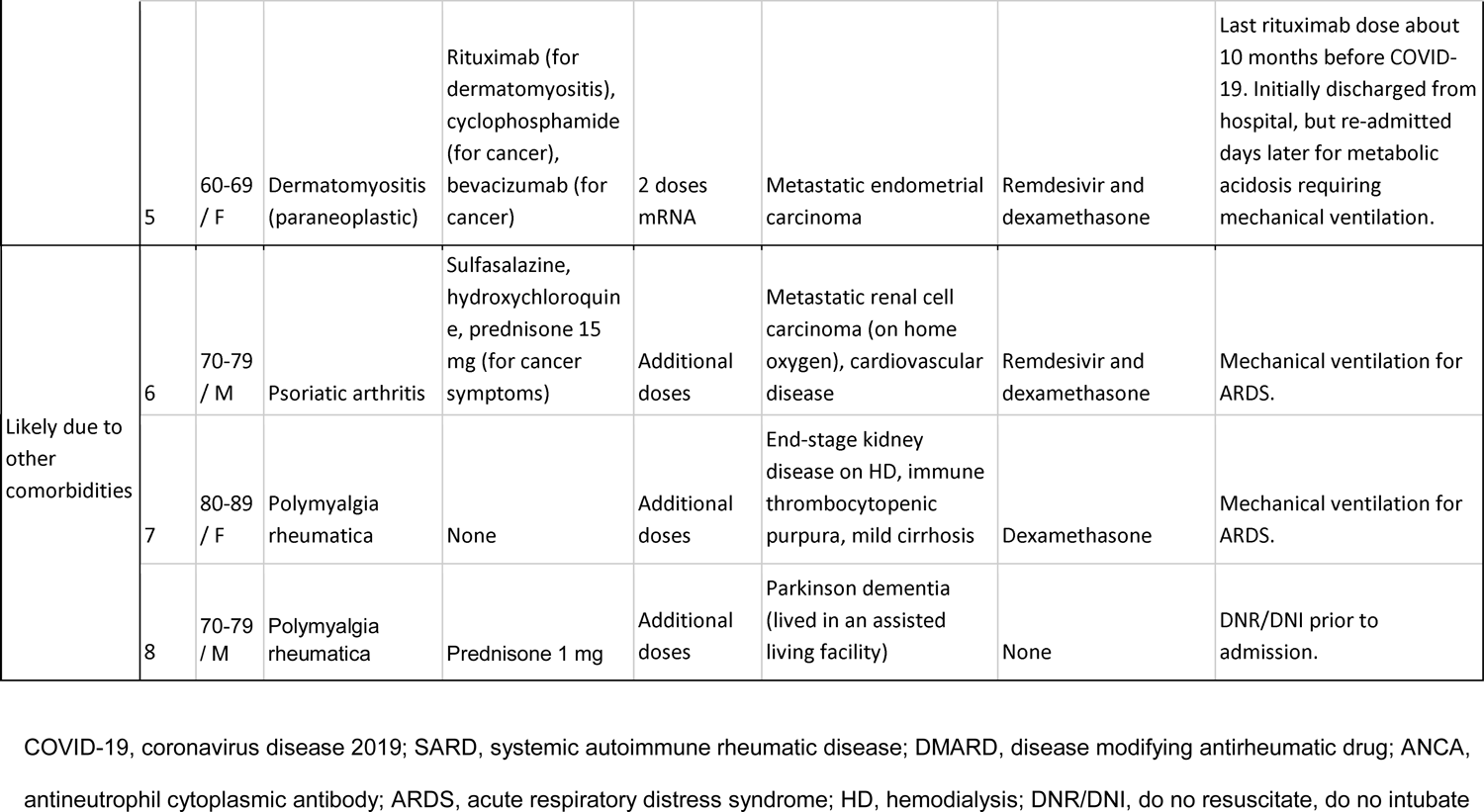
Case series of patients who died of COVID-19 during the Omicron wave

