## Supplemental files for "Temporal trends in COVID-19 outcomes among patients with systemic autoimmune rheumatic diseases: From the first wave to Omicron"

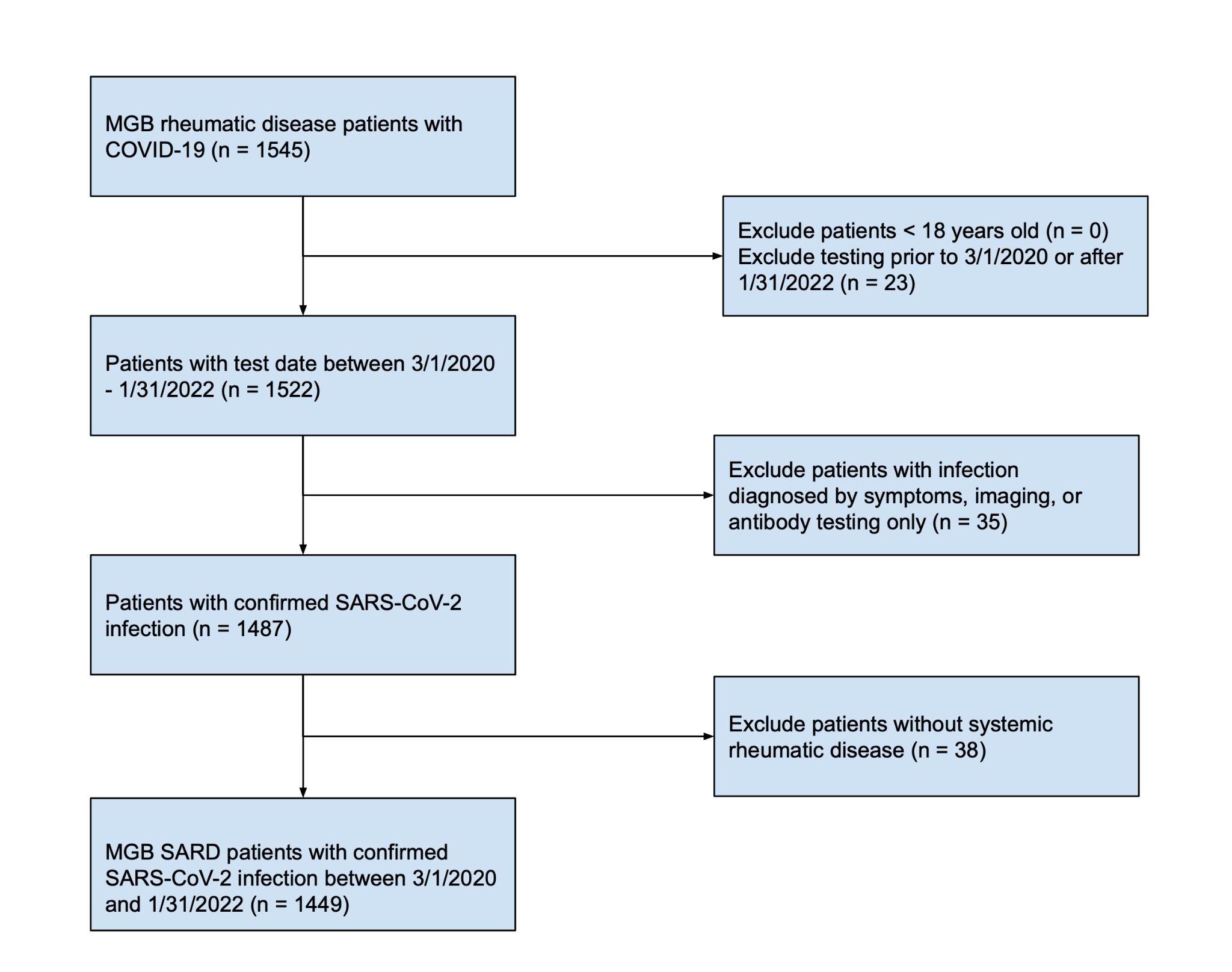

**Supplemental Figure 1:** Flow chart of patient cohort selection

MGB, Mass General Brigham; SARS-CoV-2, severe acute respiratory syndrome coronavirus 2; SARD, systemic autoimmune rheumatic disease

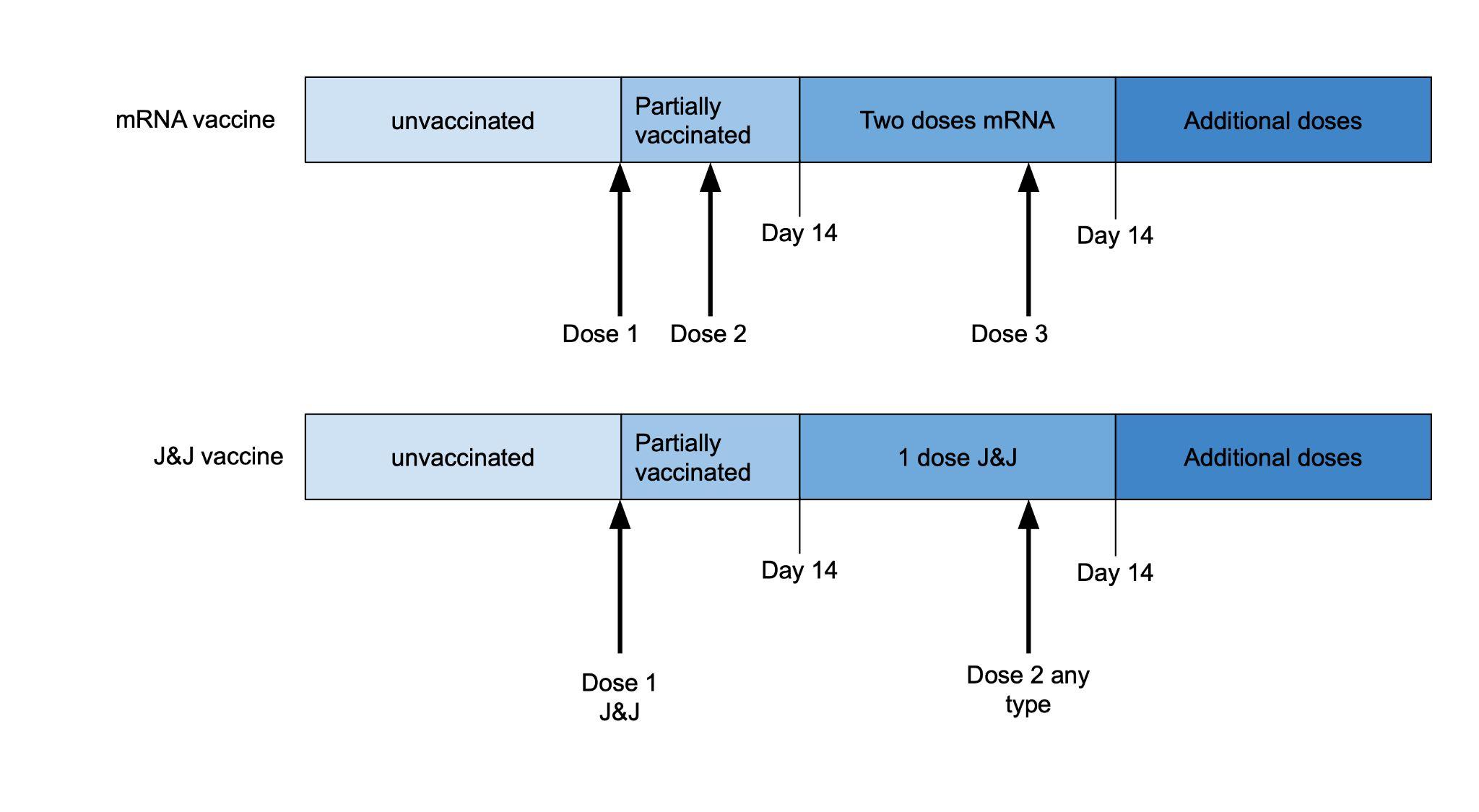

**Supplemental Figure 2:** Definition of vaccination categories: “unvaccinated,” “partially vaccinated,” “two doses mRNA or 1 dose adenovirus,” and “additional doses”

**Supplemental Table 1: Characteristics of patients with severe COVID-19 during Omicron wave**

|  | **Severe COVID-19**  **(n = 59)** | **Death (n = 8)** |
| --- | --- | --- |
| **Age, years (mean ± SD)** | 66.92 (19.1) | 77.82 (7.6) |
| **Female sex, n (%)** | 45 (76.3%) | 4 (50.0%) |
| **Race, n (%)** |  |  |
| White | 42 (71.2%) | 7 (87.5%) |
| Black | 8 (13.6%) | 1 (12.5%) |
| Asian, Hawaiian or Pacific Islander | 0 (0.0%) | 0 (0.0%) |
| Other or unknown | 9 (15.3%) | 0 (0.0%) |
| **Hispanic ethnicity, n (%)** | 3 (5.1%) | 0 (0.0%) |
| **Comorbidities, n (%)** |  |  |
| Hypertension | 38 (64.4%) | 5 (62.5%) |
| Diabetes Mellitus | 14 (23.7%) | 2 (25.0%) |
| Obesity | 19 (32.2%) | 1 (12.5%) |
| Cardiovascular disease | 16 (27.1%) | 3 (37.5%) |
| Obstructive lung disease | 14 (23.7%) | 1 (12.5%) |
| Interstitial lung disease | 6 (10.2%) | 0 (0.0%) |
| **Rheumatic disease diagnosis, n (%)** |  |  |
| Rheumatoid arthritis | 22 (37.3%) | 1 (12.5%) |
| Systemic lupus erythematosus | 5 (8.5%) | 0 (0.0%) |
| Psoriatic arthritis and spondyloarthritis | 2 (3.4%) | 1 (12.5%) |
| Other inflammatory arthritis* | 2 (3.4%) | 0 (0.0%) |
| PMR and/or GCA | 8 (13.6%) | 2 (25.0%) |
| ANCA-associated vasculitis | 8 (13.6%) | 2 (25.0%) |
| Other vasculitis** | 2 (3.4%) | 0 (0.0%) |
| Sjogren's syndrome | 3 (5.1%) | 0 (0.0%) |
| Systemic sclerosis | 1 (1.7%) | 0 (0.0%) |
| Inflammatory myopathy | 3 (5.1%) | 1 (12.5%) |
| Other connective tissue diseases† | 1 (1.7%) | 0 (0.0%) |
| Sarcoidosis | 1 (1.7%) | 0 (0.0%) |
| Multiple rheumatic diagnoses | 1 (1.7%) | 1 (12.5%) |
| Other diagnoses‡ | 0 (0.0%) | 0 (0.0%) |
| **Disease activity, n (%)** |  |  |
| Remission or low activity | 36 (61.0%) | 5 (62.5%) |
| Moderate or severe activity | 11 (18.6%) | 1 (12.5%) |
| Unknown | 12 (20.3%) | 2 (25.0%) |
| **Rheumatic disease medications at time of infection, n (%)** |  |  |
| Glucocorticoids | 28 (47.5%) | 5 (62.5%) |
| None | 31 (52.5%) | 3 (37.5%) |
| Low dose (1 to 10mg) | 20 (33.9%) | 3 (37.5%) |
| Moderate/high dose (>10 mg) | 7 (11.9%) | 2 (25.0%) |
| Unknown dose | 1 (1.7%) | 0 (0.0%) |
| Methotrexate | 11 (18.6%) | 1 (12.5%) |
| Leflunomide | 2 (3.4%) | 0 (0.0%) |
| Antimalarial | 8 (13.6%) | 2 (25.0%) |
| Sulfasalazine | 3 (5.1%) | 1 (12.5%) |
| Azathioprine | 2 (3.4%) | 0 (0.0%) |
| Mycophenolate mofetil | 5 (8.5%) | 1 (12.5%) |
| Calcineurin inhibitor | 1 (1.7%) | 1 (12.5%) |
| Cyclophosphamide | 0 (0.0%) | 0 (0.0%) |
| Belimumab | 0 (0.0%) | 0 (0.0%) |
| Rituximab | 9 (15.3%) | 2 (25.0%) |
| Abatacept | 4 (6.8%) | 1 (12.5%) |
| TNF inhibitors | 4 (6.8%) | 0 (0.0%) |
| JAK inhibitors | 2 (3.4%) | 0 (0.0%) |
| IL-6 inhibitors | 0 (0.0%) | 0 (0.0%) |
| IL-17, IL-12/23, and IL-23 inhibitors | 0 (0.0%) | 0 (0.0%) |
| IL-1 inhibitors | 1 (1.7%) | 0 (0.0%) |
| Apremilast | 0 (0.0%) | 0 (0.0%) |
| IVIG | 1 (1.7%) | 1 (12.5%) |
| **Vaccination status, n (%)** |  |  |
| Unvaccinated or pre-vaccine | 8 (13.6%) | 0 (0.0%) |
| Partially vaccinated | 6 (10.2%) | 1 (12.5%) |
| Two doses mRNA or one dose J&J | 22 (37.3%) | 1 (12.5%) |
| Additional doses | 23 (39.0%) | 6 (75.0%) |

COVID-19, coronavirus disease 2019; SD, standard deviation; PMR, polymyalgia rheumatica; GCA, giant cell arteritis; ANCA, antineutrophil cytoplasmic antibody; DMARD, disease modifying antirheumatic drug; TNF, tumor necrosis factor; IL, interleukin; JAK, Janus kinase; IVIG, intravenous immune globulin; mRNA, messenger ribonucleic acid; J&J, Johnson and Johnson / Janssen

* Includes juvenile idiopathic arthritis, other unspecified inflammatory arthritis

** Includes Takayasu's arteritis, Kawasaki disease, Behcet's disease, polyarteritis nodosa, other vasculitis

† Includes undifferentiated connective tissue disease, mixed connective tissue disease, antiphospholipid syndrome (without concurrent SLE)

‡ Includes relapsing polychondritis, IGG4 related disease, sclerosing mediastinitis, periodic fever syndromes, adult onset Still's disease

**Supplemental Table 2: Case series of patients who died of COVID-19 during the Omicron wave**

|  | **Case #** | **Age / sex** | **SARD type** | **DMARD and glucocorticoid use** | **Vaccination status** | **Notable comorbidities** | **COVID-19 treatment** | **Notes** |
| --- | --- | --- | --- | --- | --- | --- | --- | --- |
| Likely due to underlying rheumatic disease and/or immunosuppression | 1 | 70-79 / F | ANCA-associated vasculitis | Rituximab | Partially vaccinated | Inflammatory skin disease, hypertension | Monoclonal antibodies (outpatient), remdesivir and dexamethasone (inpatient) | Last rituximab infusion days before COVID-19 onset. Mechanically ventilated for ARDS. Also had pulmonary emboli. |
|  | 2 | 70-89 / F | Rheumatoid arthritis | Abatacept, hydroxychloroquine | Additional doses | Distant history of sarcoidosis, heart failure | Remdesivir and dexamethasone | Mechanical ventilation for ARDS. |
|  | 3 | 70-79 / M | ANCA-associated vasculitis | Mycophenolic acid, tacrolimus, prednisone 5 mg | Additional doses | Kidney transplant (from vasculitis) | Remdesivir and dexamethasone | Kidney transplant performed less than a year prior. Mechanical ventilation for ARDS. Also had bacterial superinfection and possible mucormycosis. |
| Possibly due to underlying rheumatic disease and/or immunosuppression | 4 | 80-89 / M | Rheumatoid arthritis | Methotrexate, prednisone 15 mg | Additional doses | Distant history of treated hepatocellular carcinoma and tonsillar squamous cell carcinoma, type 2 diabetes mellitus, heart failure, hypertension | Remdesivir and dexamethasone | Mechanical ventilation for ARDS. Also with aspiration pneumonia. |
|  | 5 | 60-69 / F | Dermatomyositis (paraneoplastic) | Rituximab (for dermatomyositis), cyclophosphamide (for cancer), bevacizumab (for cancer) | 2 doses mRNA | Metastatic endometrial carcinoma | Remdesivir and dexamethasone | Last rituximab dose about 10 months before COVID-19 onset. Initially discharged from hospital, but re-admitted days later for metabolic acidosis requiring mechanical ventilation. |
| Likely due to other comorbidities | 6 | 70-79 / M | Psoriatic arthritis | Sulfasalazine, hydroxychloroquine, prednisone 15 mg (for cancer symptoms) | Additional doses | Metastatic renal cell carcinoma (on home oxygen), cardiovascular disease | Remdesivir and dexamethasone | Mechanical ventilation for ARDS. |
|  | 7 | 80-89 / F | Polymyalgia rheumatica | None | Additional doses | End-stage kidney disease on HD, immune thrombocytopenic purpura, mild cirrhosis | Dexamethasone | Mechanical ventilation for ARDS. |
|  | 8 | 70-79 / M | Polymyalgia rheumatica | Prednisone 1 mg | Additional doses | Parkinson dementia (lived in an assisted living facility) | None | DNR/DNI prior to admission. |

COVID-19, coronavirus disease 2019; SARD, systemic autoimmune rheumatic disease; DMARD, disease modifying antirheumatic drug; ANCA, antineutrophil cytoplasmic antibody; ARDS, acute respiratory distress syndrome; HD, hemodialysis; DNR/DNI, do no resuscitate, do no intubate
